# Relation between rich-club organization versus brain functions and functional recovery after acute ischemic stroke

**DOI:** 10.1101/2020.05.26.20108563

**Authors:** Lu Wang, Kui Kai Lau, Leonard SW Li, Yuen Kwun Wong, Christina Yau, Henry KF Mak, Edward S Hui

## Abstract

Studies have shown that rich-club may underlie brain function and be associated with many brain disorders. In this study, we aimed to investigate the relation between poststroke brain functions and functional recovery versus the rich-club organization of the structural brain network of patients after first-time acute ischemic stroke. A cohort of 16 first-time acute ischemic stroke patients (11 males; 5 females) was recruited. Structural brain networks were measured using diffusion tensor imaging within 1 week and at 1, 3 and 6 months after stroke. Motor impairment was assessed using the Upper-Extremity Fugl-Meyer motor scale and activities of daily living using the Barthel Index at the same time points as MRI. The rich-club regions that were stable over the course of stroke recovery included the bilateral dorsolateral superior frontal gyri, right supplementary motor area, and left median cingulate and paracingulate gyri. The network properties that correlated with poststroke brain functions were mainly the ratio between communication cost ratio and density ratio of rich-club, feeder and local connections. The recovery of both motor functions and activities of daily living were correlated with higher normalized rich club coefficients and shorter length of local connections within a week after stroke. The communication cost ratio of feeder connections, the length of rich-club and local connections, and normalized rich club coefficients were found to be potential prognostic indicators of stroke recovery. Our results provide additional support to the notion that different types of network connections play different roles in brain functions as well as functional recovery.

## Introduction

Approximately 80% of patients suffer from motor impairment after stroke.^1,2^ Brain plasticity mechanisms, including activity-dependent rewiring and synapse strengthening, may likely underlie the recovery of brain functions.^3-6^ Previous studies have investigated the relation between motor recovery after stroke and the structural connectivity of local pathways, such as corticospinal^7-10^, alternate corticofugal^11,12^ and corticocortical pathways^13,14^. The relation between motor recovery and structural connectivity in stroke patients has also been investigated by large-scale analysis of structural connectivity^15^.

Neuroarchitecture can be understood not only at the local and global scales, as is usually performed, but also in specific groups of brain regions and the relationship among them, known as rich-club organization^16^. Rich-club organization is composed of brain regions that are closely connected, i.e., high centrality, and has been found to serve brain functions^17^. Rich-club organization may dominate the entire brain network^18^, and has been regarded as the core of communication of the whole human brain^19^. The brain regions that form rich-club organization in healthy adults are bilateral frontoparietal and subcortical regions, including the precuneus, superior frontal cortex, parietal cortex, putamen, hippocampus and thalamus.^20^ Neurological diseases and disorders, such as, schizophrenia^21^, Huntington’s disease^22^, multiple sclerosis^23^, traumatic brain injury^24^, and Alzheimer’s disease^25^, have been shown to impair rich-club organization.

Two prior studies have investigated the relation between poststroke functional outcomes related to motor function and rich-club metrics, such as the number of rich-club nodes affected by stroke^26^, and path length and the average distance between regions^27^. Considering that there is a lack of understanding of the longitudinal changes in rich-club organization after stroke, and the relation between rich-club organization versus poststroke brain functions and functional recovery, we therefore aimed to investigate whether rich-club organization would change over the course of stroke recovery, association between network metrics and poststroke brain functions, and the network metrics that may predict functional recovery.

## Materials and methods

### Subjects and functional assessments

Stroke patients (n = 16; 11 male; mean age 65.8 ± 11.0; infarct side: 50% left) with first-time acute ischemic stroke were recruited between September 2015 and July 2018 with informed consent. MRI and functional assessments were performed within 1 week (n = 12) and at 1 (n = 16), 3 (n = 13) and 6 (n = 9) months after acute ischemic stroke. Functional assessments included the Upper-Extremity Fugl-Meyer motor scale (UE-FM) and Barthel index (BI). The UE-FM was developed to quantitatively assess the severity of motor impairment of the upper extremity due to hemiplegic stroke and is based on the well-defined stages of motor recovery^28^. The BI was developed to quantitatively assess disability and functional outcomes^29^. All patients received rehabilitation at the Acute Stroke Unit of Queen Mary Hospital after admission due to acute stroke. After an average of 5 days after admission, patients were transferred to the Stroke Rehabilitation Ward of Tung Wah Hospital for more intensive rehabilitation. All patients underwent conventional occupational rehabilitation therapy sessions, including activities of daily living training, upper limb functional training, cognitive perceptual training and functional task training. All procedures were carried out following the operational guidelines of the Human Research Ethics Committee, and all protocols were approved by the Institutional Review Board of the University of Hong Kong/Hospital Authority Hong Kong West Cluster.

### Image acquisition

All MRI scans were performed using a 3.0 T MRI scanner (Achieva TX, Philips Healthcare, Best, The Netherlands) with a body coil for excitation and an 8-channel head coil for reception. Diffusion tensor imaging (DTI) was performed using single-shot spin-echo echo planar imaging, consisting of non-diffusion-weighted images (b0) and diffusion-weighted images (DWIs) with b-values of 1000 s/mm^2^ along 32 gradient directions, with the following parameters: TR/TE = 4000/81 ms, field-of-view = 230 × 230 mm^2^, reconstructed resolution = 3 × 3 mm^2^, 33 contiguous slices with thickness of 3 mm, SENSE factor = 2, number of averaging = 2, total scan time ≈ 5 minutes. TI-weighted image were acquired using 3D magnetization prepared-rapid gradient echo (MPRAGE) with the following parameters: TR/TE/TI = 7/3.17/800 ms, field-of-view = 240 × 240 mm^2^, reconstruction resolution = 1 × 1 × 1 mm^3^, 160 contiguous slices, scan time ≈ 6 min.

### Image pre-processing

All of the pre-processing procedures were performed using SPM12 (https://www.fil.ion.ucl.ac.uk/spm/). Head motion correction was performed by registering DWIs to b0 images^30^. MPRAGE images were first reoriented taking the anterior commissure^31^ as the origin. The reoriented MPRAGE images were normalized to the MNI152 template to obtain a transformation matrix *M*. The MNI152 template was then inversely normalized to MPRAGE images by the inverse of *M, M*^−1^. The inverse normalized MNI152 template was non-linearly registered to DWIs, and a transformation matrix *T* was obtained. In this way, an inverse warping transformation from the standard space to the native DTI space was obtained.

### Network construction

<u>Tractography:</u> Diffusion tensor and diffusion metrics were obtained using the Diffusion Toolkit^32^. White matter tractography was obtained using TrackVis (http://trackvis.org) using Fibre Assignment by Continuous Tracking (FACT)^33,34^ with an angle threshold of 45° and random seed of 32. Spine filter was used to smooth the fibre tracks.

<u>Network node definition:</u> The Automated Anatomical Labelling 2 (AAL2) atlas^35^ in the standard space was inversely wrapped to the individual DTI native space according to *M*^−1^ and *T*. Ninety-four cortical regions (47 in each hemisphere) were obtained, and each region was regarded as a node. Of these, 10 regions were excluded, namely, the left and right inferior occipital gyrus, left and right fusiform gyrus, left and right superior temporal pole, left and right middle temporal pole and left and right inferior temporal pole, due to variations in brain coverage.

<u>Network link definition:</u> The UCLA Multimodal Connectivity package was used to calculate the number and length of fibres between each pair of regions. Two regions were considered as connected when a fibre connected them. The link weight was defined as the fibre count between two regions.

### Network topology metrics

MATLAB (2018b) and the Brain Connectivity Toolbox (https://sites.google.com/site/bctnet/)^36^ were used to calculate brain network topology metrics. The metrics included (1) node degree, the number of nodes that a given node is connected to; (2) node strength, the sum of the weight of connected links of a node; (3) local clustering coefficient, the connection properties within the neighbourhood of a node; (4) global efficiency, the average inverse shortest path length in the network; (5) local efficiency, the global efficiency computed on node neighbourhoods; and (6) node betweenness centrality, the fraction of all shortest paths in the network that contain the node of interest.

### Rich-club organization

R (3.6.0) was used to estimate rich-club organization for each subject at each time point using a previously published method^37^. The weighted rich-club coefficient was calculated in 4 steps. First, for each node degree (k), a subnetwork was obtained by extracting the nodes with degree greater than k and the links among them. Second, for each subnetwork, the total number of links (n) and the sum of weights of the links (W) were calculated. Third, the n largest weights of the whole network were summed. Finally, the weighted rich-club coefficient of this subnetwork was subsequently calculated as follows:^38^

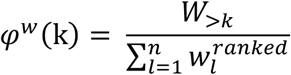

where *W_>k_* represents total weights of the links of nodes with degree larger than k, and 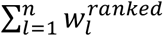 the sum of the n largest weights of the links. However, nodes with lower degree in a network have lower possibilities of sharing links with each other by coincidence, and even random networks generate increasing rich-club coefficients as a function of increasing degree threshold k. To circumvent this effect, 1,0000 random networks with the same size and same degree distribution of the network of interest were generated. The average of the rich-club coefficients of the 1,0000 random networks were calculated, *φ_random_*. The normalized rich-club coefficient of the network of interest was defined as follows:^16,37^

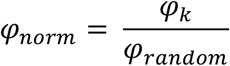

A subnetwork is considered a rich-club organization when *φ_norm_*(*k*) > 1.^21,39^

A node can be evaluated as a rich-club node when k ={k_1_, k_2_, …, k_n_}, in which the highest k was called the highest rich-club level. Each node of a rich-club organization was given a score according to their highest rich-club level Then, after averaging the score of nodes from all participants at the same time point after stroke, the top 8 nodes (i.e., 10% of all nodes) were selected as rich-club nodes.

### Node and connection types

There are two types of nodes in a brain network, namely, rich-club nodes and peripheral nodes. Furthermore, there are three types of connections in a rich-club organization (**Figure 1**), namely, rich-club connections (between rich-club nodes), feeder connections (between rich-club nodes and non-rich-club nodes), and local connections (between non-rich-club nodes). ^19^

**Figure 1.**
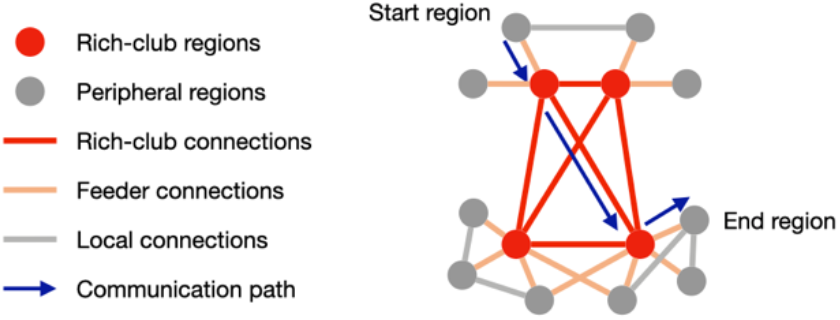
Illustration of different node and connection types. A communication path was indicated by blue arrows.

### Network communication

The cost of a link was defined as the product between its length and density (the count of streamlines between two brain regions). Communication cost was defined as the product between length and density based on their topological distance^19^. To calculate the communication cost of the three kinds of connections in a rich-club organization, first, the shortest paths between 84 nodes were calculated. Then, each link of the shortest paths was divided into three categories (rich-club, feeder and local connections). Next, the communication cost of each link was calculated by multiplying its length and density. Finally, the communication cost of three kinds of connections was obtained by summing the communication cost for links of each kind. The metric ratio of a connection type was defined as the sum of metrics of this connection type divided by the total metric of the whole network. For example, density ratio of feeder connections was defined as total density of feeder connections divided by total density of all connections. The communication cost/density ratio was defined as the communication cost ratio divided by the density ratio, which was the weight of brain communication capacity^19^.

### Statistical analysis

Statistical analyses were performed by R (version 3.6.0) with the functions lmer (https://cran.r-project.org/web/packages/lme4/lme4.pdf) and blmer (https://cran.rproject.org/web/packages/blme/blme.pdf). To simplify the statistical analyses on the local structural brain networks, the left and right hemispheres were flipped so that the left hemisphere always corresponded to the ipsilesional hemisphere. Due to attrition, some of the behavioural data and imaging data were not obtained. Imputation was thus performed on the behavioural data to increase the effective sample size for subsequent statistical analyses of prediction model. For patients no. 2, 13 and 15, the behavioural data at 6 months after stroke were imputed from those at 3 months. Since patient nos. 11 and 16 had already made full recovery at the last follow-up, full behavioural scores were assumed for the missing time points. After imputation, the 10 patients had behavioural data for all 4 time points. The imputed behavioural data are underlined in **Table 1**.

**Table 1.**
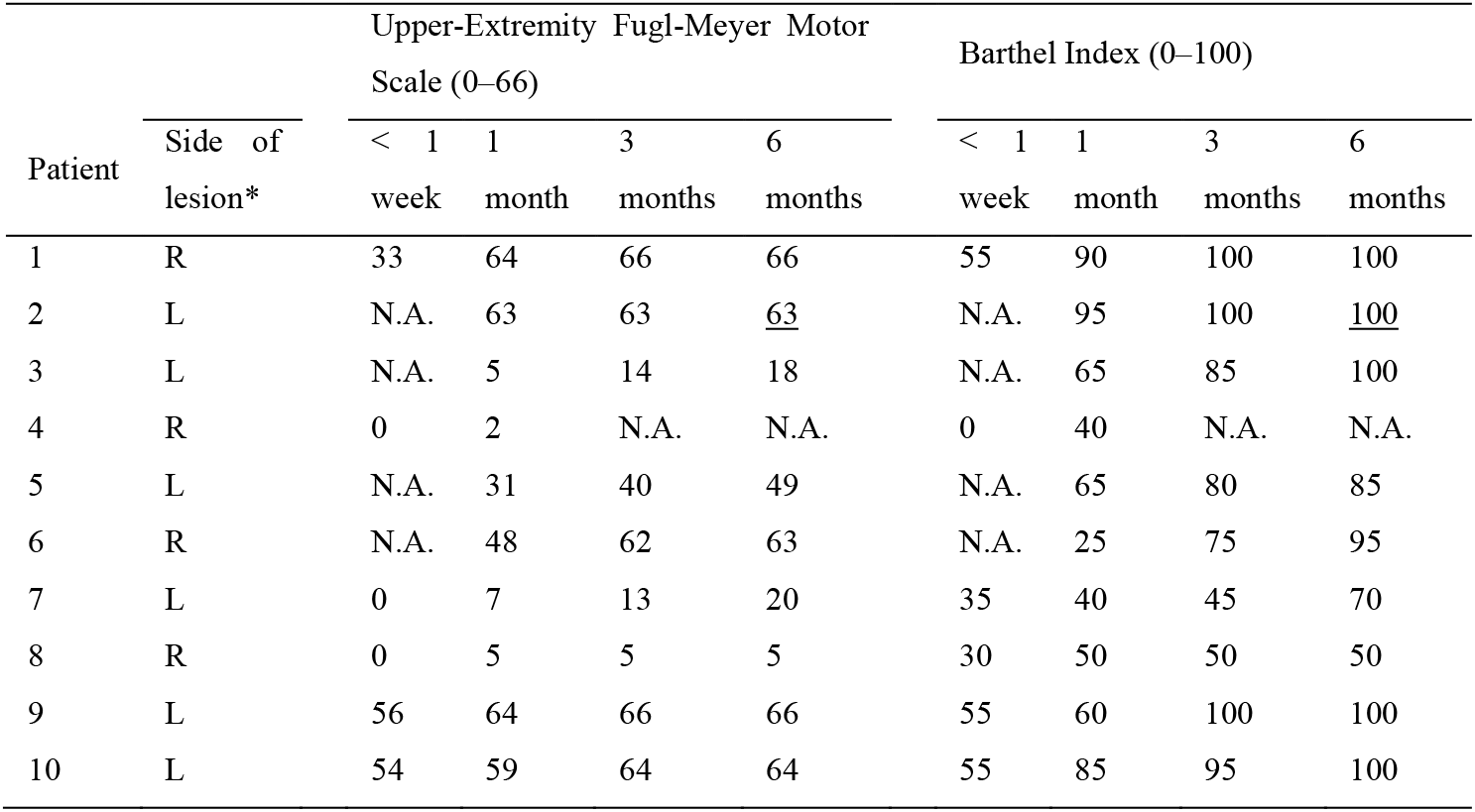
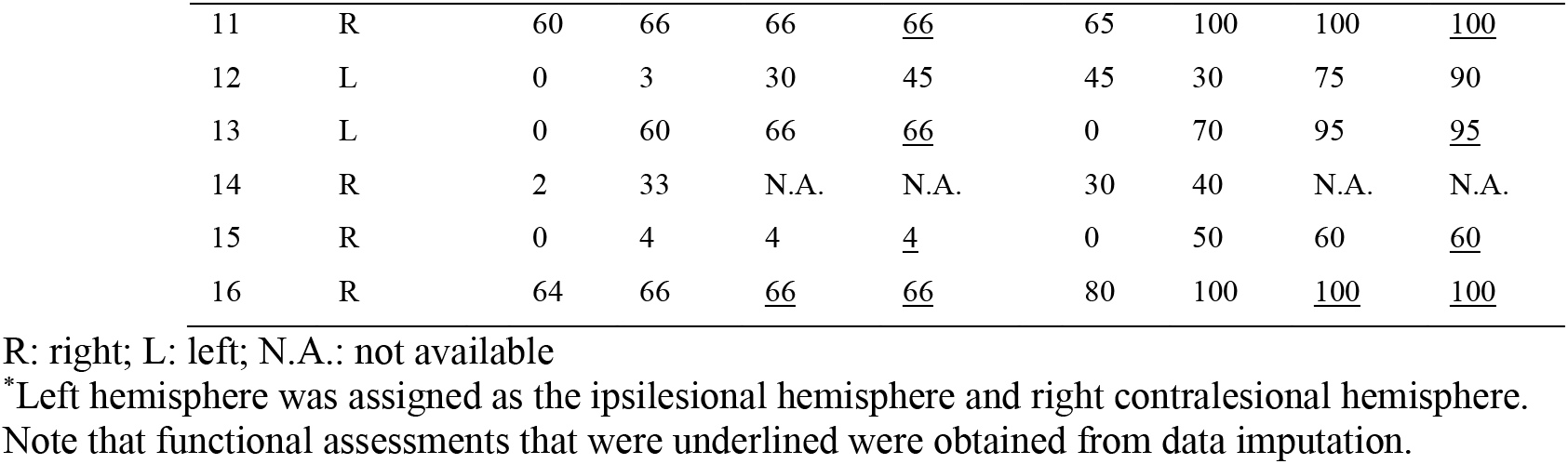
The baseline demographics and assessment of motor impairment (Upper-Extremity Fugl-Meyer motor scale) and performance in activities of daily living (Barthel Index) of n = 16 patients.

To handle data with an unequal number of longitudinal measures, a Bayesian linear mixed model^40,41^ was used as the main model in the current study so that a posteriori estimation could be maximized in a Bayesian setting. When necessary, linear mixed models were followed by post hoc tests with Bonferroni corrections for multiple comparisons at p < 0.05. This study mainly investigated three aspects: (1) to test whether functional assessments and network metrics changed with time. Models were established with the functional assessments and network metrics as responses, time as fixed variable, each subject as random variable, and age and gender as covariates. (2) To test the correlations between network metrics and functional assessments, models were established with the functional assessment data as responses, network metrics as fixed variables, each subject as random variable, and age and gender as covariates. (3) To identify biomarkers that may predict poststroke functional recovery, models were established with poststroke functional recovery (change in functional assessment from baseline) as responses, each subject as random variable, metrics at baseline as fixed variables, and time, age and gender as covariates. After selecting the metrics that may be predictors of responses, a linear mixed regression model was established by the stepwise method. In the whole study, a best fitting model was chosen by comparing models using the likelihood ratio test with the principle that the smaller the AIC was, the better the model was^42^. Additionally, the simpler model was chosen when the gap in AIC values and likelihood ratio test results was small.

## Results

### Patient demographics

MRI and functional assessments were performed on 12, 16, 13 and 9 patients within 1 week and at 1, 3 and 6 months after first-time acute ischemic stroke, respectively, as shown in **Figure 2**. A linear mixed model was used to test the fixed effect of time on functional assessments. UE-FM *(β_time_* = 3.47; 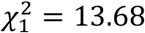, p < 0.001) and BI *(β_time_* = 7.94, 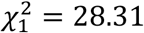, p < 0.001) significantly increased with time. Post hoc tests showed that UE-FM scores at 1 (p = 0.01), 3 (p < 0.001) and 6 (p < 0.001) months were significantly higher than those within 1 week after stroke. BI scores at 1, 3 and 6 (p < 0.001, all) months were significantly higher than those within 1 week after stroke, and BI scores at 3 and 6 (p < 0.001, all) months were significantly higher than 1 month after stroke.

**Figure 2.**
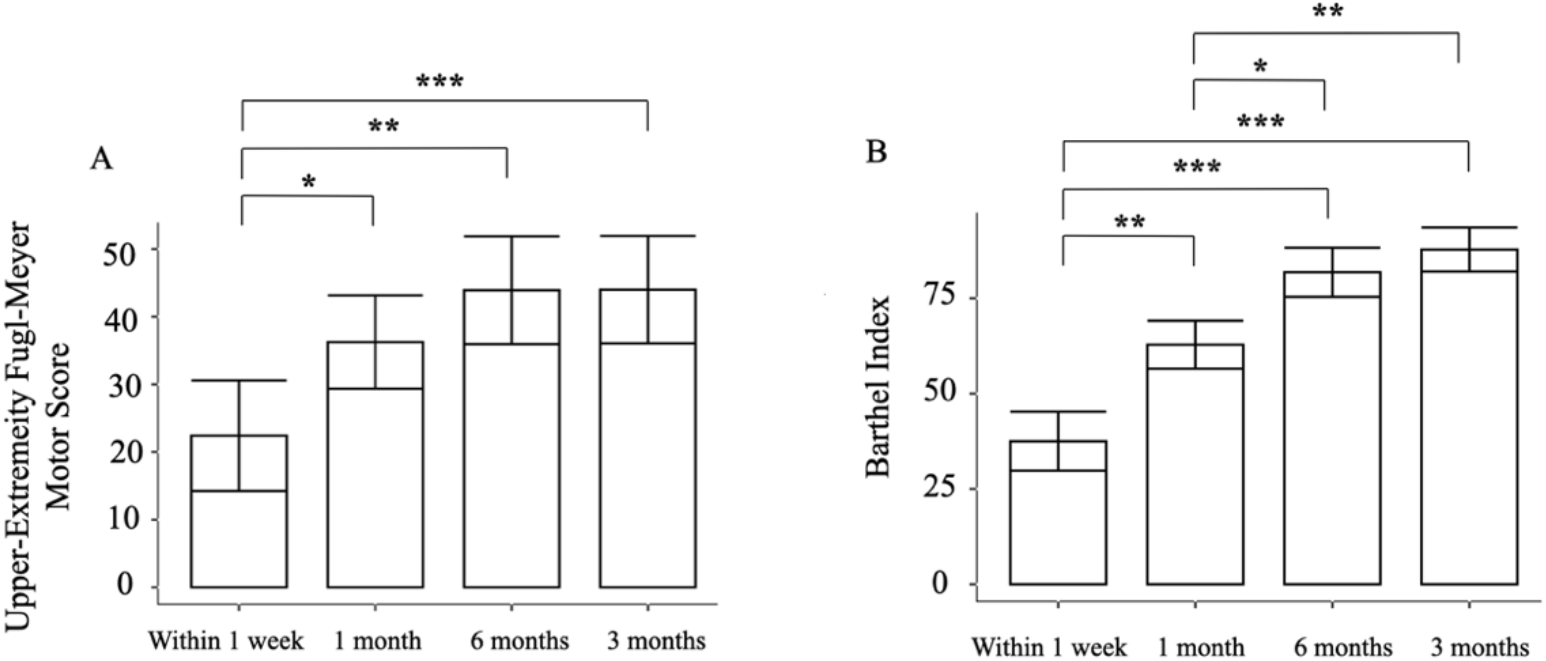
Bar chart with error bars (standard deviations) of the (A) Upper-Extremity Fugl-Meyer motor scale and (B) Barthel index scores within 1 week (n = 12), and at 1 (n = 16), 3 (n = 13) and 6 (n = 9) months after first-time acute ischemic stroke. Post hoc tests with Bonferroni corrections for multiple comparisons across different time points were performed (*p <0.05, **p<0.01, ***p<0.001).

### Rich-club organizational changes after stroke

Rich-club organization was found from the structural brain network of patients within 1 week, and at 1, 3 and 6 months after first-time acute ischemic stroke (**Figure 3**). Not every patient (grey lines) had a normalized rich-club coefficient larger than 1 except at 6 months after stroke. Linear mixed model was used to test the effect of time after stroke on normalized rich-club coefficients with age and gender as covariates. Significant time effect on the normalized rich-club coefficients at **k** = 20 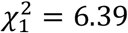, *p* = 0.011, *β_time_* = 0.16), 21 (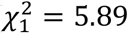, *p* = 0.015, *β_time_* = 0.15), k = 22 (*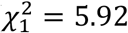*, *p* = 0.015, *β_time_* = 0.16), and k = 23 (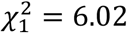, *p* = 0.014, *β_time_* = 0.16) were observed. The brain regions that were rich-club regions at each time point after stroke were listed in **Table 2** and shown in **Figure 4**. Notice that bilateral dorsolateral superior frontal gyrus, right supplementary motor area, and left median cingulate and paracingulate gyri remained as rich-club regions at all 4 time points.

**Figure 3.**
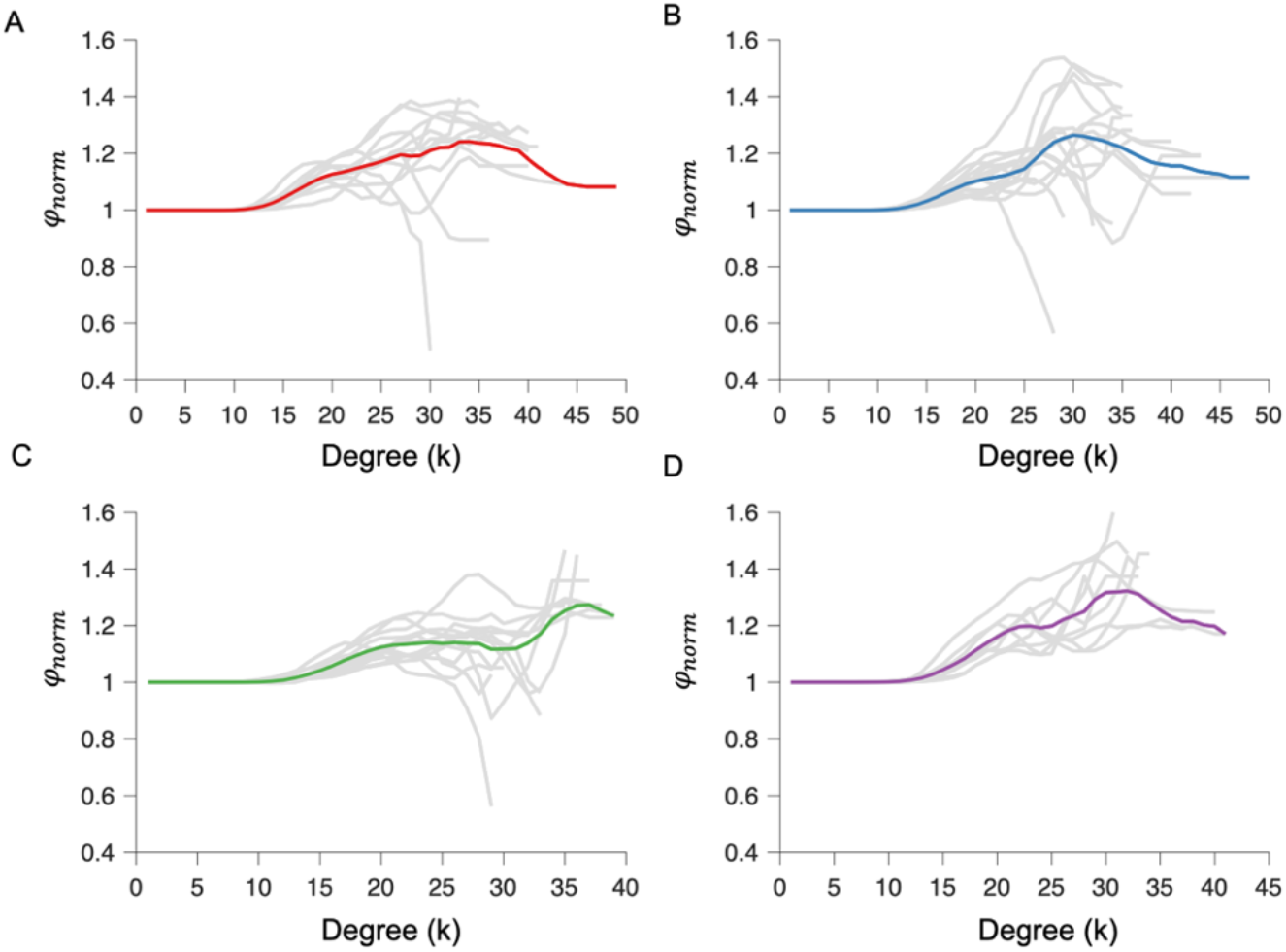
Normalized rich-club coefficient of the structural brain network of patient within (A) 1 week and at (B) 1, (C) 3 and (D) 6 months after first-time acute ischemic stroke. Grey lines: individual patient; Colored lines: average of all patients.

**Table 2.** Rich-club regions at each time point after first-time acute ischemic stroke.

| Rich-club regions | Side* | Time after acute ischemic stroke |  |  |  |
| --- | --- | --- | --- | --- | --- |
|  |  | < 1 week | 1 month | 3 months | 6 months |
| Dorsolateral superior frontal gyrus | Left | √ | √ | √ | √ |
|  | Right | √ | √ | √ | √ |
| Supplementary motor area | Left | √ | √ |  |  |
|  | Right | √ | √ | √ | √ |
| Insular | Left | √ |  | √ | √ |
|  | Right | √ | √ |  |  |
| Anterior cingulate and paracingulate gyri | Left |  | √ | √ | √ |
|  | Right | √ |  |  |  |
| Median cingulate and paracingulate gyri | Left | √ | √ | √ | √ |
|  | Right |  | √ | √ | √ |
| Precuneus | Right |  |  | √ |  |
| Putamen | Left |  |  |  | √ |

\*Left hemisphere was assigned as the ipsilesional hemisphere and right contralesional hemisphere.

**Figure 4.**
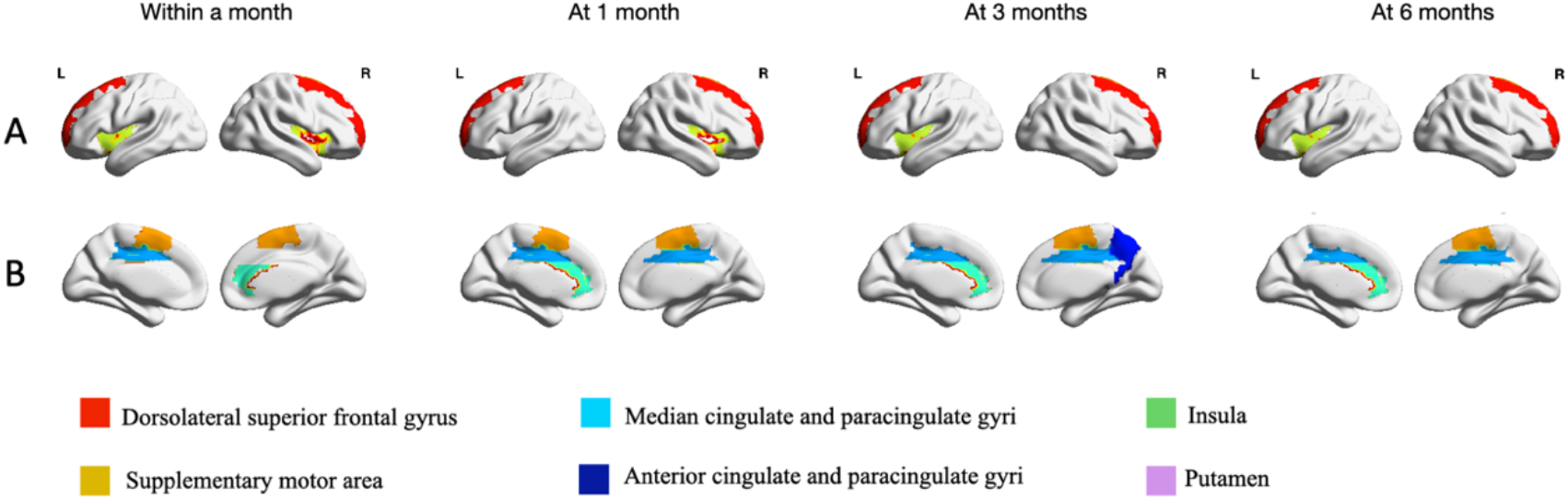
Rich-club regions at each time point after first-time acute ischemic stroke.

### Longitudinal changes in network metrics after first-time acute ischemic stroke

The structural brain networks at local and rich-club scales over the course of stroke recovery were examined. Metrics of the network at local scale pertain to the network metrics of each brain region; rich-club scale to the mean link metrics of rich-club, feeder and local connections. A linear mixed model was used to test the fixed effect of time after stroke on these network metrics with age and gender as covariates, and subject as random variable. For the local scale, the node degree, node strength, local clustering coefficient, local efficiency and nodal betweenness centrality of a number of nodes changed with time after stroke (**Figure 5**). For the rich-club scale (**Table 3**), significant negative time effect on the length of feeder connections (*β_time_* = −0.13, 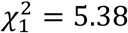, *p* = 0.020) and communication cost ratio/density ratio of local connections (*β_time_* = −0.16, 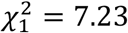, *p* = 0.007) were observed. Significant positive time effect on the density ratio of rich club connections (*β_time_* = 0.15, 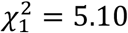, *p* = 0.024) and communication cost ratio/density ratio of feeder connections (*β_time_* = 0.13, 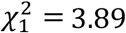, *p* = 0.049) were observed.

**Figure 5.**
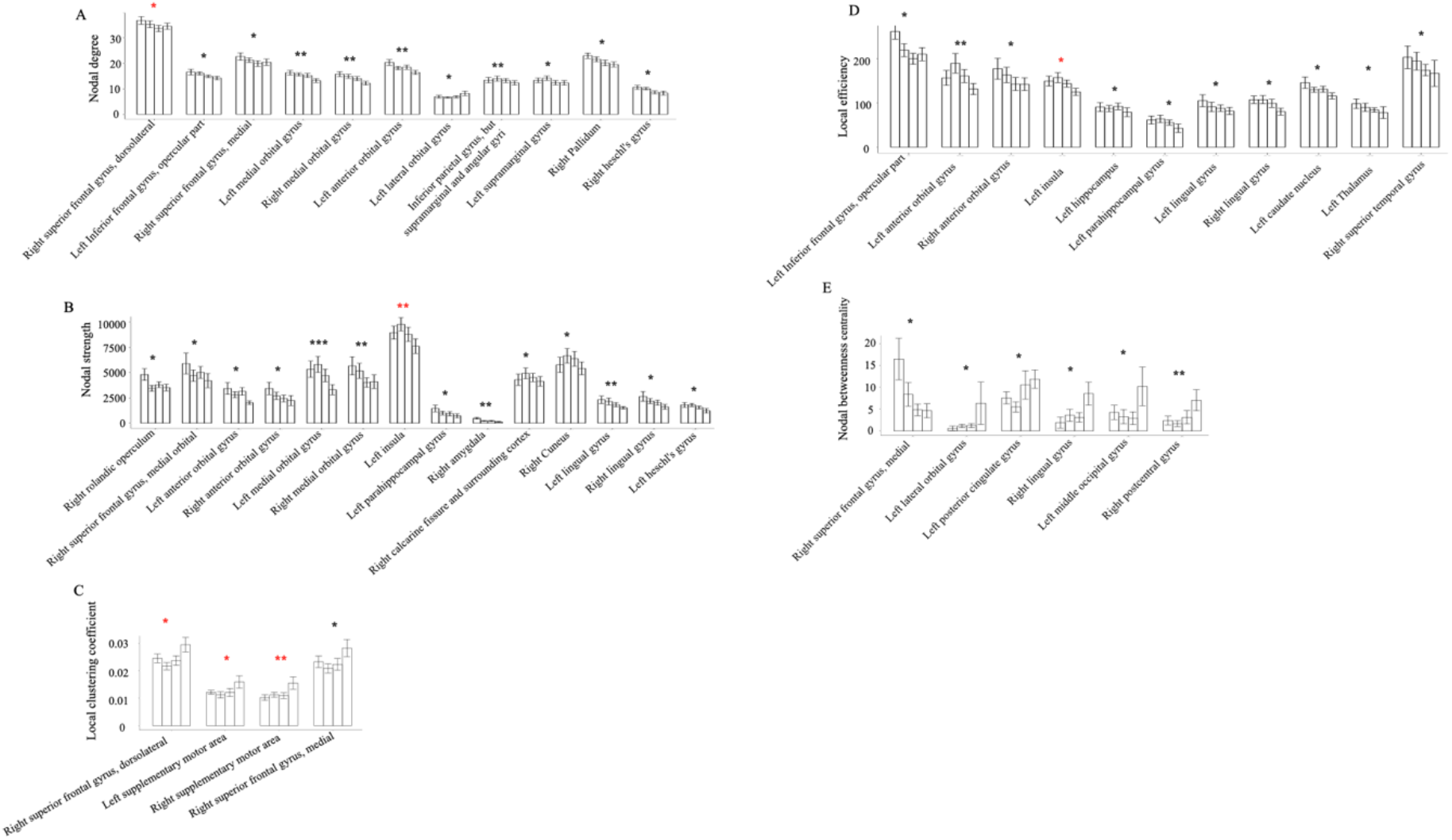
The effect of time after first-time acute ischemic stroke on the metrics of brain network at local scale: (A) node degree, (B) node strength, (C) local clustering coefficient, (D) local efficiency and (E) nodal betweenness centrality. Only regions showing statistical significance were displayed. **\***p<0.05, **\*\***p<0.01, **\*\*\***p<0.001. Rich-club regions were annotated with a red **\***.

**Table 3.** The effect of time after first-time acute ischemic stroke on the metrics of brain network at rich-club scale. Only metrics showing statistical significance were shown.

| Connection Type | Network metric | $\beta_0$ | $\beta_{time}$ | $\chi^2$ | Df | Pr (> $\chi^2$) |
| --- | --- | --- | --- | --- | --- | --- |
| **Rich-club** | Density ratio | 0.18 | 0.15 | 5.10 | 1 | 0.024 |
|  | Length | 0.06 | -0.13 | 5.38 | 1 | 0.020 |
| **Feeder** | Communication cost ratio/density ratio | -1.30 | 0.13 | 3.89 | 1 | 0.049 |
| **Local** | Communication cost ratio/density ratio | 2.60 | -0.16 | 7.23 | 1 | 0.007 |

$\beta_0$ and $\beta_{\text{time}}$ are the fixed effects coefficients: $\beta_0$ is the intercept, $\beta_{\text{time}}$ is the linear slope; Df: degrees of freedom for the chi-square test.

### Relation between poststroke brain functions and network metrics

A linear mixed model was used to test the relation between functional assessments and the metrics of brain network at rich-club scale with age and gender as covariates, and subject as random variable. The length (*β*_1_ = −6.09, 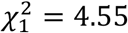, *p* = 0.033) and cost (*β*_1_ = −6.77, *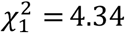*, *p* = 0.037) of feeder connections, and the communication cost ratio/density ratio of rich club conections (*β*_1_ = 4.54, 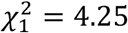, *p* = 0.039) were correlated with UE-FM. The communication cost ratio/density ratio of local (*β*_1_ = −10.05, 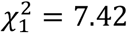, *p* = 0.006), feeder (*β*_1_ = 7.47, 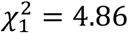, *p* = 0.028), rich-club connections (*β*_1_ = 11.60, 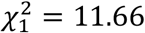, *p* = 0.001), and the density ratio of rich-club connections (*β*_1_ = 9.24, 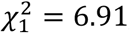, *p* = 0.009) were correlated with BI. All statistical results were summarized in **Table 4**.

**Table 4.**
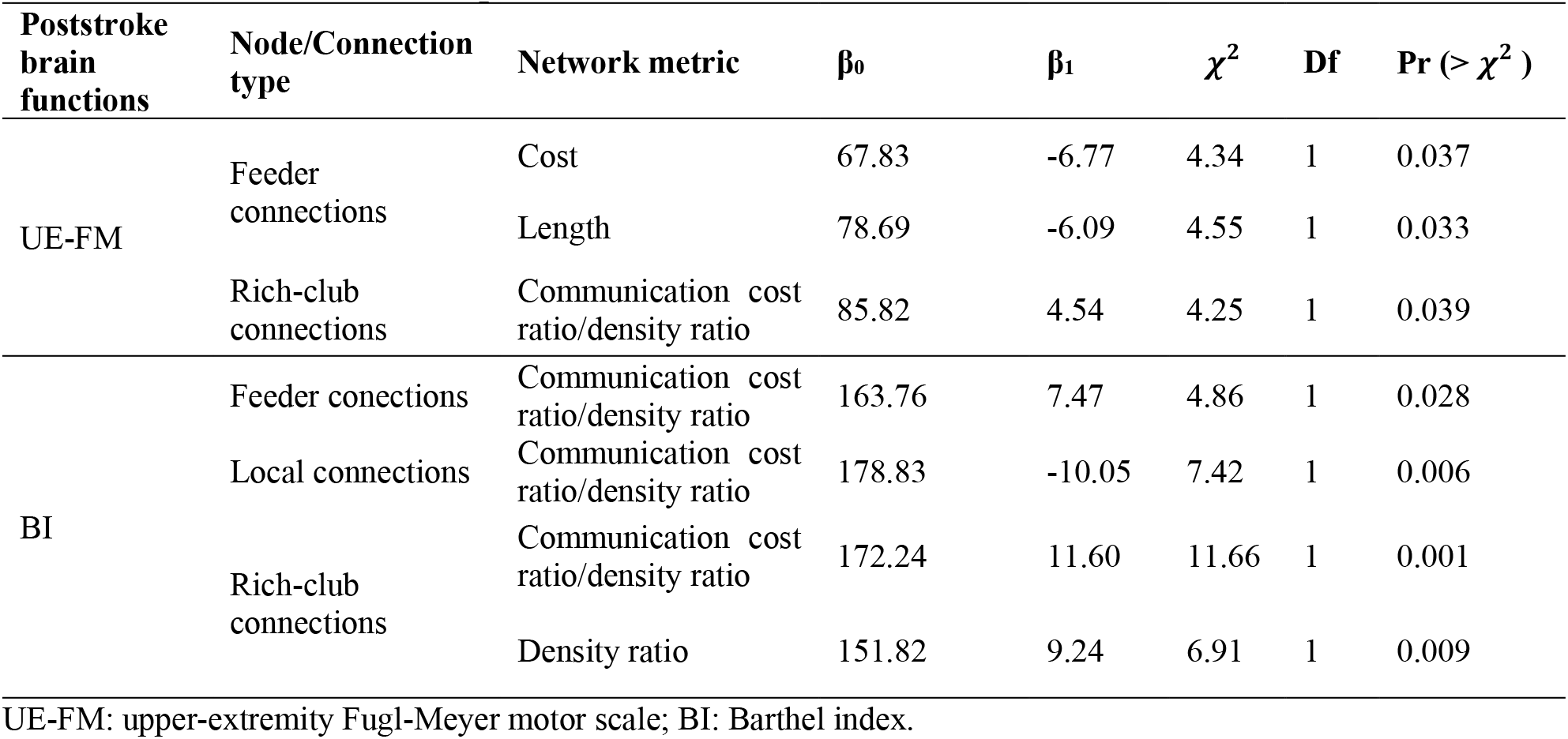
Correlation between poststroke brain functions versus rich-club metrics.

| Poststroke brain functions | Node/Connection type | Network metric | $\beta_0$ | $\beta_1$ | $\chi^2$ | Df | Pr ($> \chi^2$) |
| --- | --- | --- | --- | --- | --- | --- | --- |
| UE-FM | Feeder connections | Cost | 67.83 | -6.77 | 4.34 | 1 | 0.037 |
|  |  | Length | 78.69 | -6.09 | 4.55 | 1 | 0.033 |
|  | Rich-club connections | Communication cost ratio/density ratio | 85.82 | 4.54 | 4.25 | 1 | 0.039 |
| BI | Feeder connections | Communication cost ratio/density ratio | 163.76 | 7.47 | 4.86 | 1 | 0.028 |
|  | Local connections | Communication cost ratio/density ratio | 178.83 | -10.05 | 7.42 | 1 | 0.006 |
|  | Rich-club connections | Communication cost ratio/density ratio | 172.24 | 11.60 | 11.66 | 1 | 0.001 |
|  |  | Density ratio | 151.82 | 9.24 | 6.91 | 1 | 0.009 |

UE-FM: upper-extremity Fugl-Meyer motor scale; BI: Barthel index.

### Prediction of poststroke functional recovery using metrics of brain network at rich-club scale

Linear mixed models were used to investigate the relation between changes in poststroke brain functions from baseline (i.e., poststroke functional recovery) and baseline metrics of brain network at rich-club scale as well as normalized rich-club coefficient. For the change in UE-FM, the length of rich-club connections (*β*_1_ = 8.42, 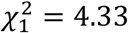, *p* = 0.037), normalized rich-club coefficients at k = 19 (*β*_1_ = 14.06, 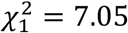, *p* = 0.008), k = 20 (*β*_1_ = 11.06, 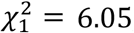, *p* = 0.014), the density ratio (*β*_1_ = 9.49, 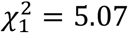, *p* = 0.024), cost ratio (*β*_1_ =9.29, 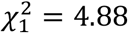, *p* = 0.027), communication cost ratio (*β*_1_ = 21.59, 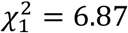, *p* = 0.009) of feeder connections, and the length (*β*_1_ = −12.96, 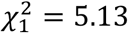, *p* = 0.024), density ratio (*β*_1_ = −8.94, 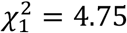, *p* = 0.029), cost ratio (*β*_1_ = −9.50, *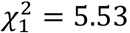*, *p* = 0.015) and communication cost ratio/density ratio (*β*_1_ = −12.42, *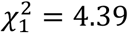*, *p* = 0.036) of local connections were significantly correlated. For the change in BI, the density (*β*_1_ = −12.62, 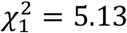, *p* = 0.024), length (*β*_1_ = −12.74, 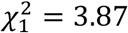, *p* = 0.049) and cost (*β*_1_ = −13.18, 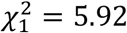, *p* = 0.015) of local connections, and normalized rich-club coefficients were significantly correlated. These results were summarized in **Table 5**.

**Table 5.**
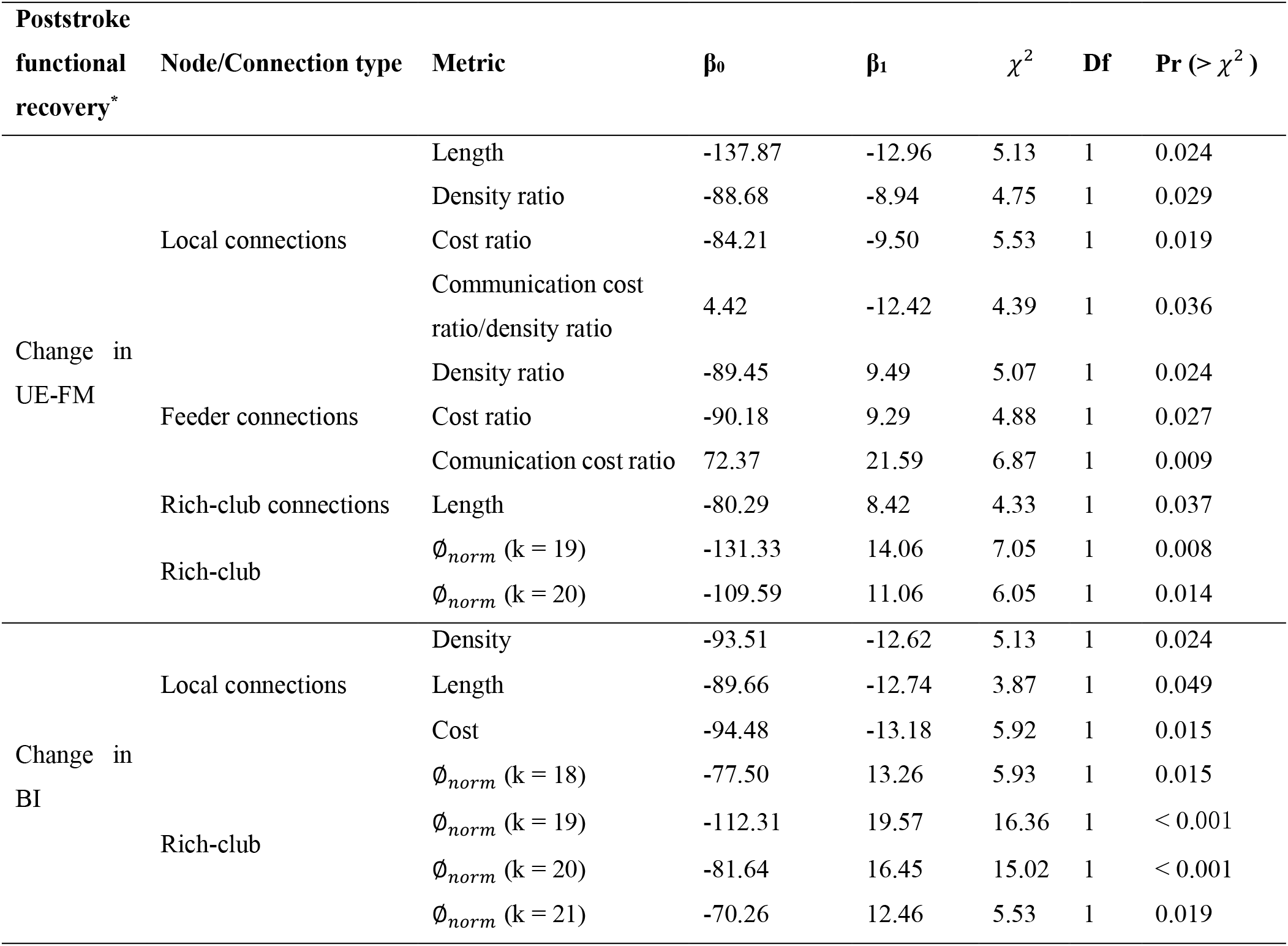
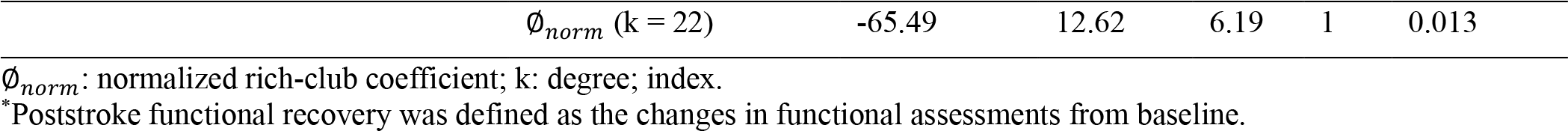
Correlation between poststroke functional recovery versus metrics of brain network at rich-club scale.

The network metrics that were selected to predict poststroke functional recovery were shown in **Table 6**. The communication cost ratio of feeder connections, length of rich-club and local connections, and normalized rich-club coefficient (k = 19) could predict UE-FM changes (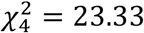, p < 0.001). The length of local connections and normalized rich-club coefficients (k =18, 19) could predict BI changes (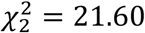, p < 0.001).

**Table 6.** Summary of prediction models.

| Poststroke functional recovery | Model | Variables | AIC | BIC | $\chi^2$ | Df | Pr ($> \chi^2$) |
| --- | --- | --- | --- | --- | --- | --- | --- |
| Change in UE-FM | Baseline | Age, gender, time | 192.67 | 200.88 |  |  |  |
| | Prediction | Age, gender, time, communication cost ratio of feeder connections, length of rich-club and local connections, $\Phi_{norm}$ (k = 19) | 177.35 | 191.02 | 23.33 | 4 | < 0.001 |
| Change in BI | Baseline | Age, gender, time | 244.39 | 252.59 |  |  |  |
| | Prediction | Age, gender, time, length of local connections, $\Phi_{norm}$ (k = 18, 19) | 228.79 | 241.09 | 21.60 | 2 | < 0.001 |

$\Phi_{norm}$ : normalized rich-club coefficient; k: degree; index.
AIC: Akaike information criterion; BIC: Bayesian information criterion.

## Discussion

Our study aims to investigate the relation between poststroke brain functions and functional recovery versus the rich-club organization of the structural brain network of patient after first-time acute ischemic stroke. First of all, we investigated the effect of time after stroke on brain functions, rich-club organization and metrics of the brain network at rich-club and local scales. Second, we examined the relation between poststroke brain functions versus rich-club organization and network metrics. Finally, we established models to predict poststroke functional recovery using the metrics of brain network at rich-club scale and normalized rich-club coefficient measured at baseline.

### Longitudinal changes in brain network after acute ischemic stroke

The rich-club regions that remained unchanged over the course of stroke recovery included the bilateral dorsolateral superior frontal gyri, right supplementary motor area, and left median cingulate and paracingulate gyri **(Table 2, Figure 4**). As much as half of the rich-club regions changed during stroke recovery may suggest that recovery may require remodelling of a significant portion of rich-club organization. It is noteworthy that the rich-club regions of the patients at 6 months after stroke, who have largely functionally recovered, were not the same as those in normal controls as reported by van den Heuvel et al^20^. This discrepancy may suggest that their networks may be different, although effects due to the difference in postprocessing could not be ruled out.

The properties of the local network of a number of brain regions, majority of which were not rich-club regions, changed with time after stroke (**Figure 4**). For network connections, the the density ratio of rich-club connections increased with time after stroke (**Table 3**). Considering that rich-club regions are highly connected brain regions and density ratio reflects the how strong the connection is, our results suggest that this increase may be an attempt of the rich-club organization to further improve communication efficiency to aid poststroke functional recovery^19^. For feeder and local connections, their density ratio manifested contrasting time effect, likely suggesting a rebalance of resources, thereby allowing higher communication capacity for feeder connections that connect rich-club to non-rich-club regions. Schirmer and Chung showed in a study of the rich-club organization across the life span that the communication path of feeder connections strengthened, and that of local connections weakened^43^. Together, these results suggest that rich-club and non-rich-club regions may likely play very different roles over time.

### Plausible neuroarchitectural underpinning of poststroke brain functions and functional recovery

Our results showed that of all the network properties that correlated with poststroke functions, communication cost ratio/density ratio of rich-club connections positively correlated with both motor function and daily activities of daily living (**Table 4**). Considering that the benefit of rich-club organization is to confer short communication relays to the brain network as a whole, albeit its high cost of wiring, our results likely suggest that poststroke brain functions may hinge on the communication efficiency conferred by the rich-club organization^19^.

For the neuroarchitectural underpinning of poststroke functional recovery (change in functional assessment from baseline), our results showed that higher normalized rich club coefficients within a week after stroke were correlated with the recovery of both motor functions and activities of daily living (**Table 5**), suggesting that residual rich-club organization of structural brain network after acute stroke may play an important role in supporting poststroke functional recovery. For the recovery of motor functions, it is interesting to note that it was correlated with lower density ratio and cost ratio of local connections, higher density ratio and cost ratio of feeder connections. Our results provide additional support to the notion that rich-club and non-rich-club regions likely play very different roles not only in aging^43^ but also stroke recovery. For the recovery of activities of daily living, it was negatively correlated with the density, cost and length of local connctions, indicating that local connections may play a more dominant role on the recovery of activities of daily living.

Of the baseline network metrics that correlate with poststroke functional recovery, the communication cost ratio of feeder connections, length of rich-club connections, length of local connections, and normalized rich-club coefficient could potentially be prognostic indicators of stroke recovery (**Table 6**). Our findings extended the work by Ktena et al ^27^ and Schirmer et al ^26^ in modelling poststroke functional outcome. Both of their work demonstrated that the prediction of stroke functional outcome could be improved by the incorporation of the number of rich-club regions that were affected by stroke.

### Limitations

Our study still has several limitations. First, the infarct was located in different hemispheres among the 16 patients. To avoid systematic error, we swapped the brain regions from the two hemispheres for patients with right-sided infarct to ensure that all infarcts were on the left hemisphere. However, brain asymmetry may have confounded our results; for example, the strengths of the left and right hemispheres are not equal^39^. Second, there were some missing data in our dataset due to patient attrition. We have nonetheless partially circumvented this problem using data imputation. Finally, we did not have data from control group in order to investigate the effect of stroke on rich-club organizations, and to compare the difference in rich-club organization between functionally recovered stroke patients versus healthy controls.

## Conclusion

We have successfully demonstrated the relation between poststroke brain functions and functional recovery versus rich-club organization and network metrics after first-time acute ischemic stroke. Our results provide additional support to the notion that different types of network connections play different roles in brain functions as well as functional recovery.

## Data Availability

Data available on request due to privacy/ethical restrictions

## Data availability

Available upon request.

## Funding

This work was supported by the Research Grants Council of the Hong Kong Special Administrative Region, China, HKU 17108514.

## Conflict of interest

None.

## Ethics approval

This study was approved by the local Institutional Review Board

## Patient consent

Informed consent was obtained from all patients.

## Permission to reproduce material from other sources

Not applicable.

## Clinical trial registration

Not applicable.

## Notes

### Competing Interest Statement

The authors have declared no competing interest.

### Funding Statement

This work was supported by the SK Yee medical foundation and Research Grants Council of the Hong Kong Special Administrative Region, China, HKU 17108514.

### Author Declarations

Human Research Ethics Committee

